# Natural History and Acute-phase Predictors of Medium Coronary Artery Aneurysm Outcomes in Kawasaki Disease: A 3-year Prospective Cohort Study

**DOI:** 10.1101/2025.08.22.25334272

**Authors:** Ying Liu, Shuran Shao, Yimin Hua, Kaiyu Zhou, Jinlin Wu, Chuan Wang

## Abstract

**Background:** Kawasaki disease (KD), the leading cause of pediatric acquired coronary artery aneurysms (CAAs), exhibits heterogeneous outcomes in medium CAAs (4–8 mm), which lack standardized risk stratification. This study aimed to identify acute-phase predictors of prognostic outcomes in medium CAAs to optimize monitoring strategies.

**Methods:** A single-center retrospective cohort analyzed 78 KD patients with acute-phase medium CAAs (2013–2021). Serial echocardiography assessed CAA evolution, defining persistence (medium/giant CAAs) and progression (≥8% diameter increase). Multivariate logistic regression, ROC analysis, and Kaplan-Meier survival curves evaluated associations between clinical/laboratory parameters and outcomes.

**Results:** The persistent group (n=23) exhibited significantly prolonged prothrombin time (PT: 15.69 ± 5.53 vs. 12.68 ± 2.18 s, *P* = 0.014) and larger maximum CAA diameters (maximal CAA: 6.69 ± 1.35 vs. 5.04 ± 0.97mm, *P*<0.001). Multivariate analysis identified PT (OR=1.47, 95% CI:1.04–2.08, *P* = 0.031) and maximal CAA (OR=3.29, 95% CI:1.16–9.35, *P* = 0.025) as independent predictors of persistence. ROC-derived thresholds for risk stratification included PT >13.6s (AUC=0.712) and maximal CAA ≥5.65mm (AUC=0.857). Kaplan-Meier analysis confirmed significant divergence in persistence between threshold-stratified groups (log-rank P<0.001 for maximal CAA).

**Conclusion:** Acute-phase PT elevation and maximal CAA diameter ≥5.65mm are robust predictors of medium CAA persistence. These findings advocate for intensified surveillance in high-risk subgroups, integrating coagulation profiles and serial echocardiography to mitigate long-term coronary complications.

## Introduction

Kawasaki Disease (KD) is an acute systemic vasculitis that predominantly affects children under the age of five^1^. Despite the administration of intravenous immunoglobulin (IVIG) together with aspirin has greatly reduced the risk of coronary involvements, 20-30% of KD patients develop transient coronary artery dilatation (CAD) and 5-9% will develop coronary artery aneurysms (CAAs) during the acute stage^2, 3^. The management and prognosis of KD differ significantly based on the duration and magnitude of CAAs.

The prognosis for CAD or small CAAs is generally favorable, with most patients experiencing complete resolution without long-term complications^4–6^. In contrast, patients with large CAAs mostly show a possible persistence and often require close monitoring as well as aggressive anti-coagulation therapy or interventional procedures^5–13^. Patients with medium CAAs, however, present a more complex and heterogeneous clinical course. These aneurysms may stabilize, regress partially or progress to larger ones or stenotic lesions due to intimal thickening and fibrosis in the chronic stage^14–16^.

Despite the clinical significance of medium CAAs, their natural history remains poorly characterized, and predictive factors for their different prognostic outcomes are inadequately understood. Previous studies have often grouped all CAAs together^4, 17^, obscuring the unique prognostic features and predictive factors specific to medium CAAs. Furthermore, the predictive value of acute-phase indicators, such as inflammatory markers and echocardiographic findings, has not been thoroughly investigated in this subset of patients^5, 18^.

This prospective study aims to address these gaps by focusing exclusively on medium CAAs in a cohort of KD patients receiving standardized treatment. Through systematic follow-up using echocardiography, we seek to delineate the natural history of medium CAAs and identify acute-phase predictive factors associated with their different prognosis. By doing so, this study will provide critical insights into early risk stratification and inform tailored management strategies for children with medium CAAs, ultimately improving long-term outcomes.

## Methods

### Study design

Patients diagnosed with KD who received IVIG treatment within 10 days from fever onset were prospectively enrolled from January 2013 and December 2021 at the West China Second University Hospital. The diagnosis of KD is based on criteria established by the American Heart Association’s scientific statement^2^ of KD and a pediatric cardiologist and experienced pediatric echocardiographers determined the diagnosis and measured CAAs, respectively. Exclusion criteria primarily consisted of other connective tissue diseases such as systemic lupus erythematosus (SLE), as well as other conditions known to cause coronary artery aneurysms, including congenital coronary artery fistula and chronic active Epstein-Barr virus (EBV) infection, et cetera. In addition, for patients enrolled during and after the COVID-19 pandemic, SARS-CoV-2 infection was excluded through PCR testing.

### Treatment

All patients received the same standard treatment^2^ for KD. High-dose IVIG (2 g/kg given as a single intravenous infusion) and aspirin (30-50 mg/kg/day) were initially administered within 10 days of illness onset. Initial IVIG resistance was defined as recurrent or persistent fever or other clinical signs of KD for at least 36 h but not longer than 7 days after initial IVIG treatment. For patients with IVIG resistance, the second IVIG (2g/kg given as a single intravenous infusion) was given according to the expert consensus for the diagnosis and treatment for KD in China. Furthermore, pulse intravenous methylprednisolone (10-30mg/Kg/day for 3 consecutive days) followed by oral prednisone (2mg/Kg/day) tapered for 7 days were applied as the additional treatment if the patient had recurrent or persistent fever even after the second IVIG administration. Antithrombotic therapy was applied according to CAA size as follows: Patients without aneurysms received low-dose aspirin (3-5 mg/kg/day) for 6-8 weeks. Those with small CAAs were maintained on low-dose aspirin monotherapy until aneurysm resolution was documented. For medium CAAs, dual antiplatelet therapy (aspirin 3-5 mg/kg/day plus clopidogrel 0.2 mg/kg/day) was administered. Patients with giant CAAs received combined anticoagulation (warfarin with target INR 2.0-2.5) plus low-dose aspirin, with periodic monitoring of anticoagulation intensity.

### Data Collection

Demographic, clinical, and laboratory data were systematically extracted from electronic medical records and structured questionnaires. Clinical parameters encompassed days of illness before admission, IVIG resistance, use of methylprednisolone and diagnostic criteria for incomplete KD and recurrent KD. Principal KD symptoms recorded were bilateral non-purulent conjunctivitis, perioral edema, rash, cervical lymphadenopathy (≥1.5 cm), and extremity erythema/edema.

Laboratory biomarkers collected during the acute phase (pre-IVIG) and 48 hours after initial IVIG (post-IVIG), included complete blood count (CBC), coagulation profile, liver and renal function tests, sodium, potassium, lipid profile, C-reactive protein (CRP), and erythrocyte sedimentation rate (ESR). Derived indicators such as neutrophil-to-lymphocyte ratio (NLR), platelet-to-lymphocyte ratio (PLR), lymphocyte-to-CRP ratio (LCR) and systemic immune-inflammation index (SII) were calculated.

### Echocardiographic evaluation and follow-Up

All echocardiograms were performed by a single senior echocardiologist using a EPIQ7 ultrasound system (Philips Healthcare, Andover, USA). The absolute dimensions of the left main coronary artery (LMCA), left anterior descending artery (LAD), left circumflex (LCX) and right coronary artery (RCA) were recorded from the parasternal short-axis images. Measurements were obtained starting at LAD and LCX openings, extending to their junction with LMCA. Additional measurements were taken 0.2–0.5 cm distal to RCA, LAD, and LCX origins. When CAAs occurred in multiple branches, largest aneurysm diameter was recorded and corresponding Z-scores calculated. CAAs were defined using Japanese Circulation Society criteria^1^: luminal diameter >3 mm in children <5 years or >4 mm in older children, or 1.5× adjacent segment diameter. The CAAs were classified as small CAAs (localized dilatation with <4 mm internal diameter or 2.5 ≤ Z-score <5), medium CAAs (aneurysms with an internal diameter from ≥4 to <8 mm or 5≤Z-score <10), and giant CAAs (aneurysms with an internal diameter of ≥8 mm or 10≤Z-score). In this study, the size of the coronary artery and maximal CAA was defined as the largest one during the first month after KD onset. Besides, the number and location of CAAs had also been included for further analysis.

A total of 79 KD patients with medium CAA during the first month after disease onset were enrolled. After excluding 1 case lost during follow-up, 78 patients were included in the final analysis. Follow-up commenced at hospital discharge, with all enrolled patients undergoing scheduled echocardiographic evaluations at 2 weeks, 1 month, 2 months, 6 months, 1 year, 2 years, and 3 years post-discharge, as most CAAs that undergo regression typically regress within 3 years when spontaneous resolution occurs.

Patients were stratified into two groups based on the follow-up results, including regressed group (n=55) and persistence group (n=23) (Figure 1). Regressed group was defined as normalization of diameter or existence of small CAAs, confirmed with the use of computed tomography or angiography if indicated when the results of a 2-dimensional echocardiogram showed regression of medium-sized coronary aneurysms, while persistent group included patients with medium and giant CAAs. Progression^19^ was defined as ≥8% diameter increment across three consecutive echocardiograms since the second month. Informed written consent was obtained from the parents after the nature of the study had been fully explained to them. The University Ethics Committee on Human Subjects at Sichuan University approved the study.

**Figure 1.**
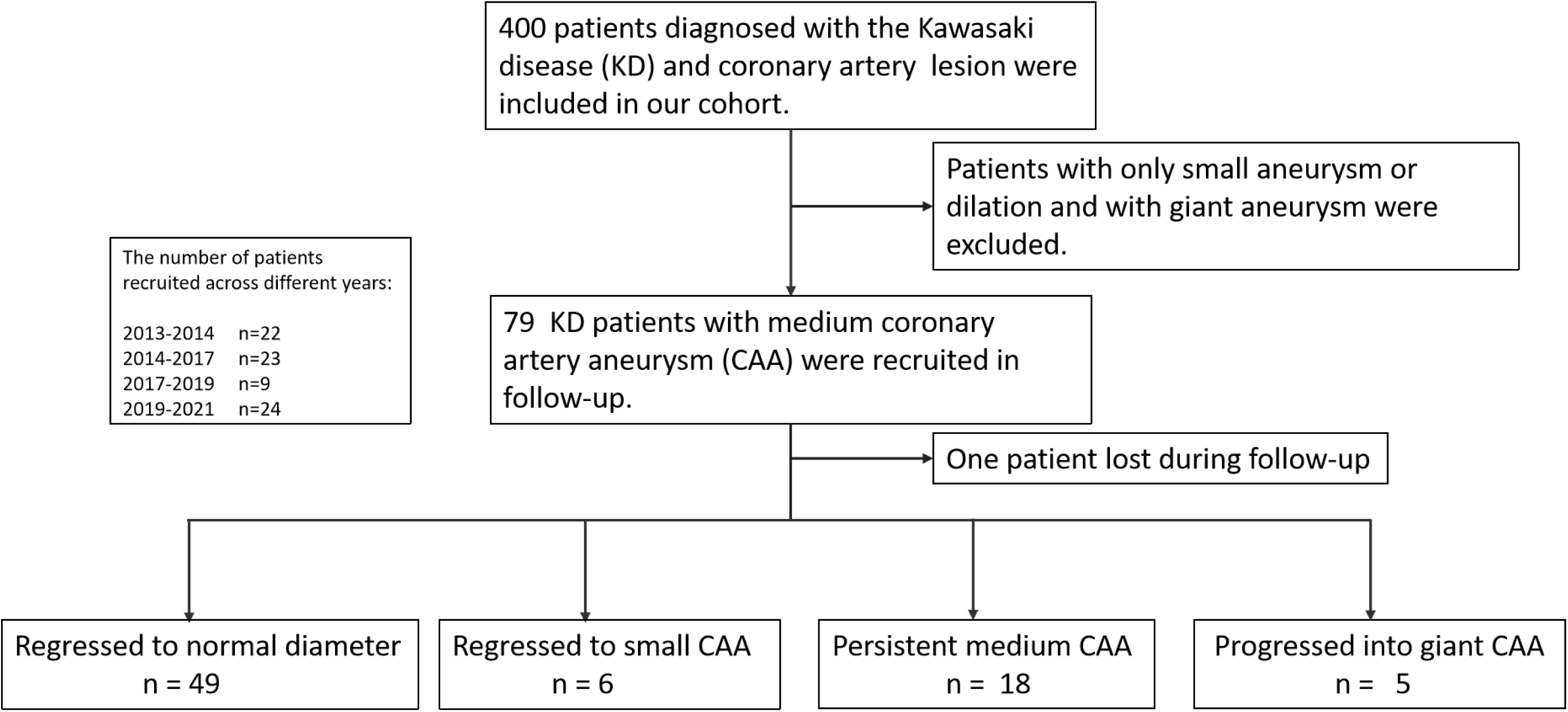
Flow chart of patients with Kawasaki disease (KD) and medium coronary artery aneurysm (CAA). Initially, 400 KD patients with coronary artery lesion were recruited in our cohort. After excluding patients with only artery dilation or small CAA, or with both medium and giant CAA, 79 patients proceeded to the follow-up process. Expect that one patient was lost to follow-up at the 1-month interval, the remaining patients continued follow-up until 3 years.

### Statistical Analysis

Statistical analyses were conducted as follows: Initial assessment of data normality and homogeneity was performed using Shapiro-Wilk and homogeneity of variance tests. Based on these results, quantitative variables were summarized as mean±standard deviation for parametric data, whereas qualitative variables were expressed as frequencies (percentages). For between-group comparisons, continuous variables were analyzed using either Student’s t-test or Mann-Whitney U test, while categorical variables were evaluated through χ² test or Fisher’s exact test as appropriate. To delineate independent predictors of CAA persistence, we first implemented multivariate logistic regression adjusted for relevant covariates. Subsequently, receiver operating characteristic (ROC) curve analysis was utilized to establish optimal diagnostic thresholds for key predictors. Temporal patterns of CAA regression were comparatively analyzed using Kaplan-Meier survival curves with log-rank testing. All analyses were executed in EmpowerStats (v3.3), with statistical significance defined as a two-tailed P < 0.05.

## Results

### Study population

78 KD patients with medium CAAs during acute period were included in final analysis. Based on the final echocardiographic assessments, 55 patients were classified into the regressed group, among whom 49 achieved normalized coronary artery diameters and 6 retained small CAAs. Conversely, 18 patients exhibited persistent medium CAAs, while 5 demonstrated progressive enlargement of CAAs during follow-up, ultimately progressing to giant CAAs. The proportion of medium CAAs regressing to small or normal size differed significantly across follow-up periods: 56% within the first year, compared to only 10% in the second year, and merely 4% in the third year. Specifically, among medium CAAs that regressed to small CAA or normal size within 3 years, the majority (78%) of these regressions occurred within the first year. Furthermore, progression from medium to giant CAA size occurred exclusively within the first year with a percentage of 6.4%. In addition, no thrombosis, coronary stenosis, acute myocardial infarction or death was documented (Figure 2).

**Figure 2.**
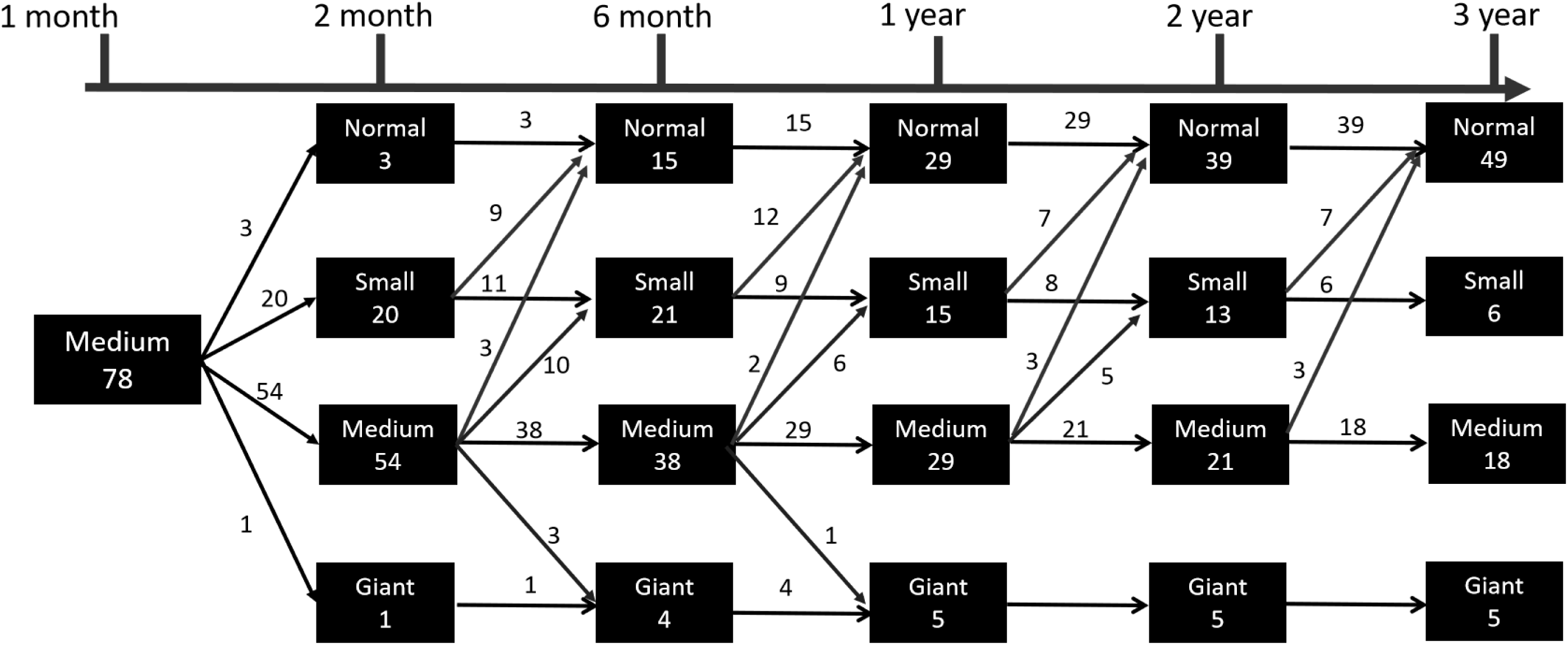
The coronary artery aneurysm status of Kawasaki disease patients at different time points during follow-up.

### Risk factors associated with CAA persistence

Although no significant differences were observed in baseline demographics, clinical manifestations, initial and repeated IVIG resistant rates between groups, the persistent group exhibited a significantly higher incidence of recurrent KD compared to the regression group (14.3% vs. 1.8%, P=0.028). Analysis of pre-IVIG laboratory parameters revealed elevated neutrophil percentages in the persistent group (66.6%±19.7 vs. 76.3%±14.2, P=0.038), whereas other inflammatory markers, hepatic and renal function indices, electrolyte levels (sodium, potassium), and lipid profiles showed no significant difference. Notably, the persistent group demonstrated prolonged prothrombin time (PT) values (12.7±2.2s vs. 15.7±5.5s, P=0.014). Significant disparities in coronary artery morphology were also observed between the two groups. The persistent CAA group exhibited significantly larger RCA diameters at initial (4.5±1.3mm vs. 6.0±2.0mm, P=0.001) and maximal CAA diameters at 1 month (5.0 ± 1.0mm vs. 6.7 ± 1.3mm, P<0.001), along with higher prevalence of multiple CAAs (P =0.043) and bilateral involvement (38.6% vs. 66.7%, P=0.027). Furthermore, aneurysmal progression occurred more frequently in this group (17.5% vs. 85.7%, P <0.001) (Table 1). In addition, post-IVIG laboratory parameters remained comparable between groups (Supplementary table 1).

**Table 1.**
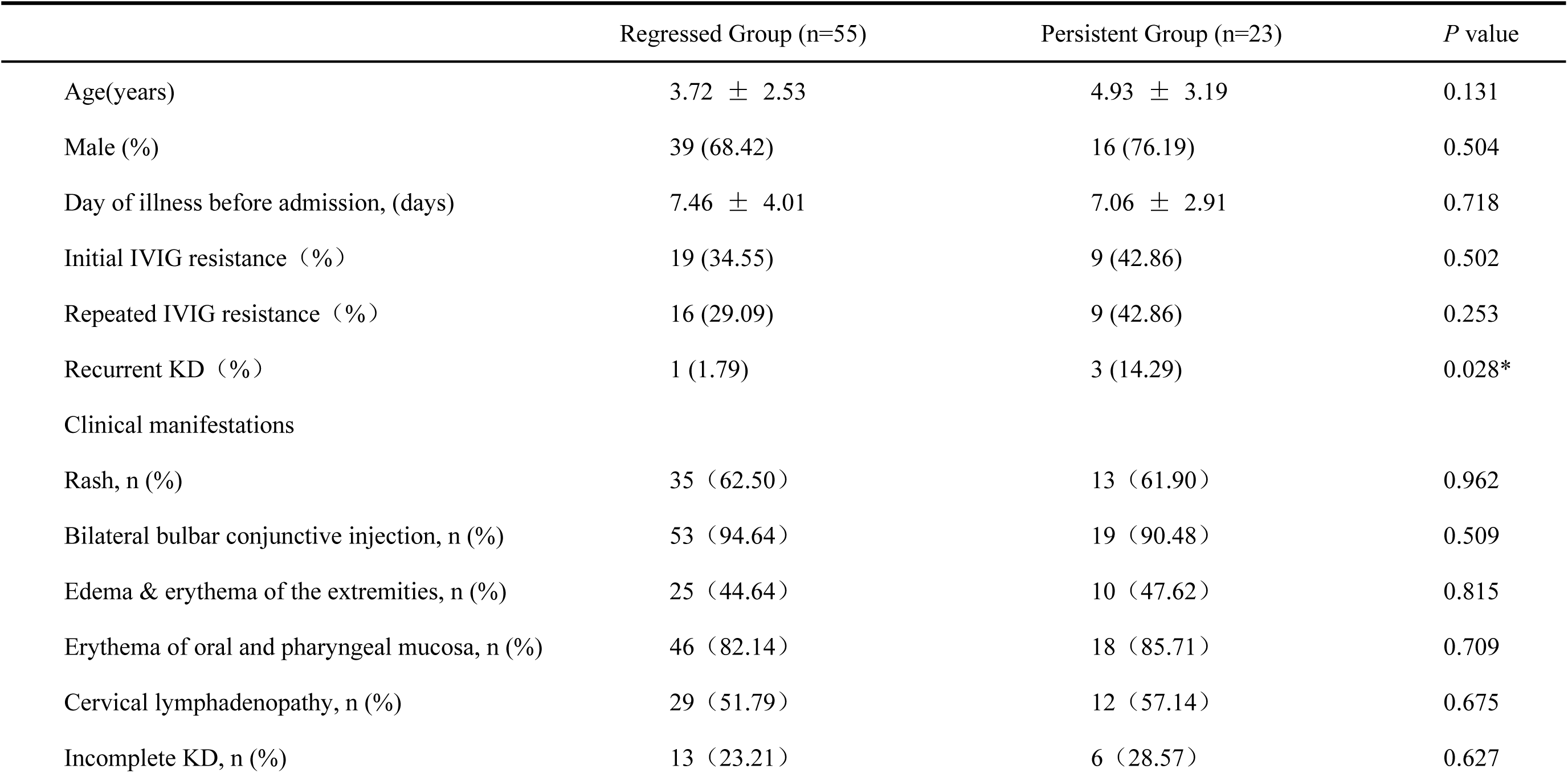
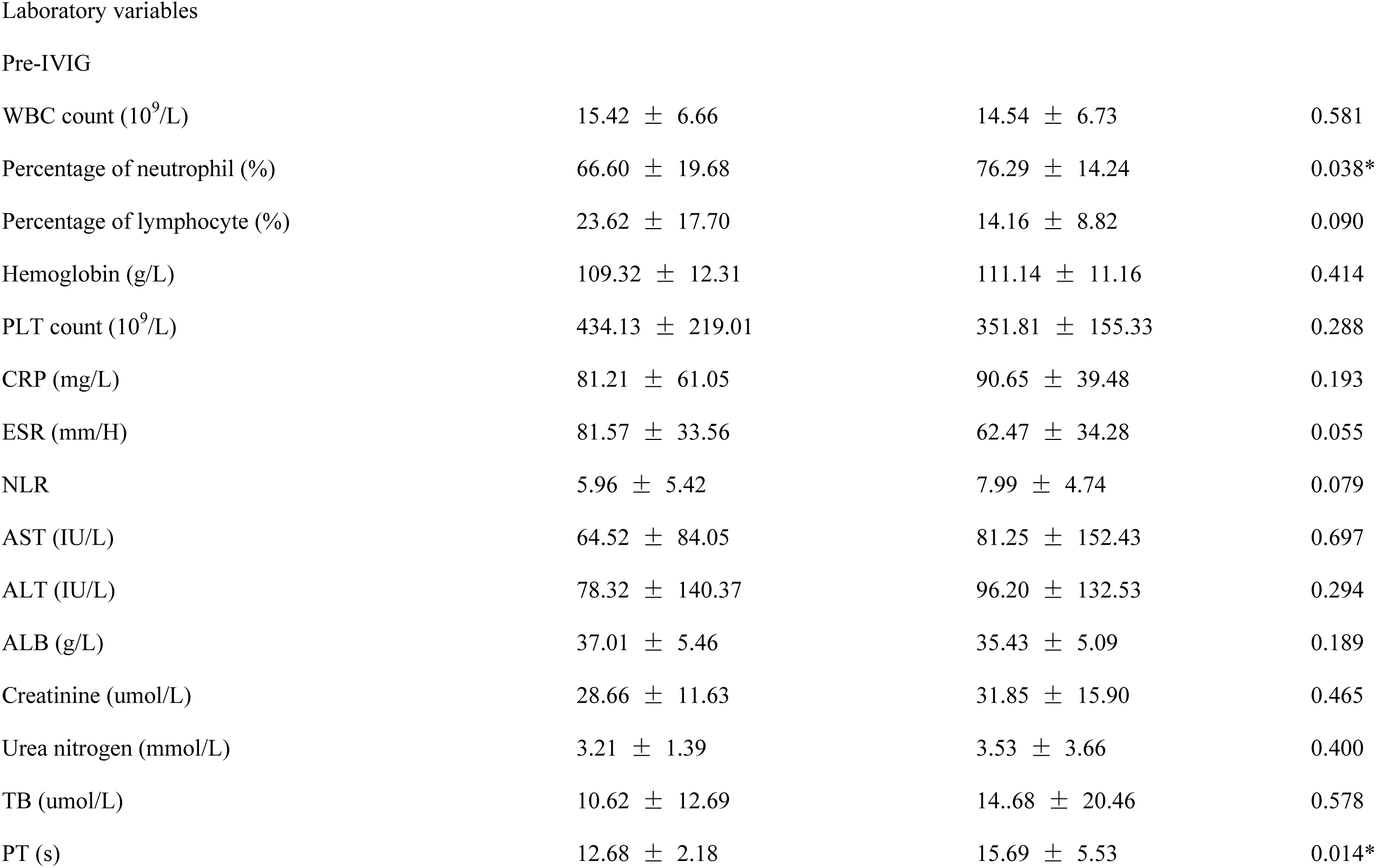
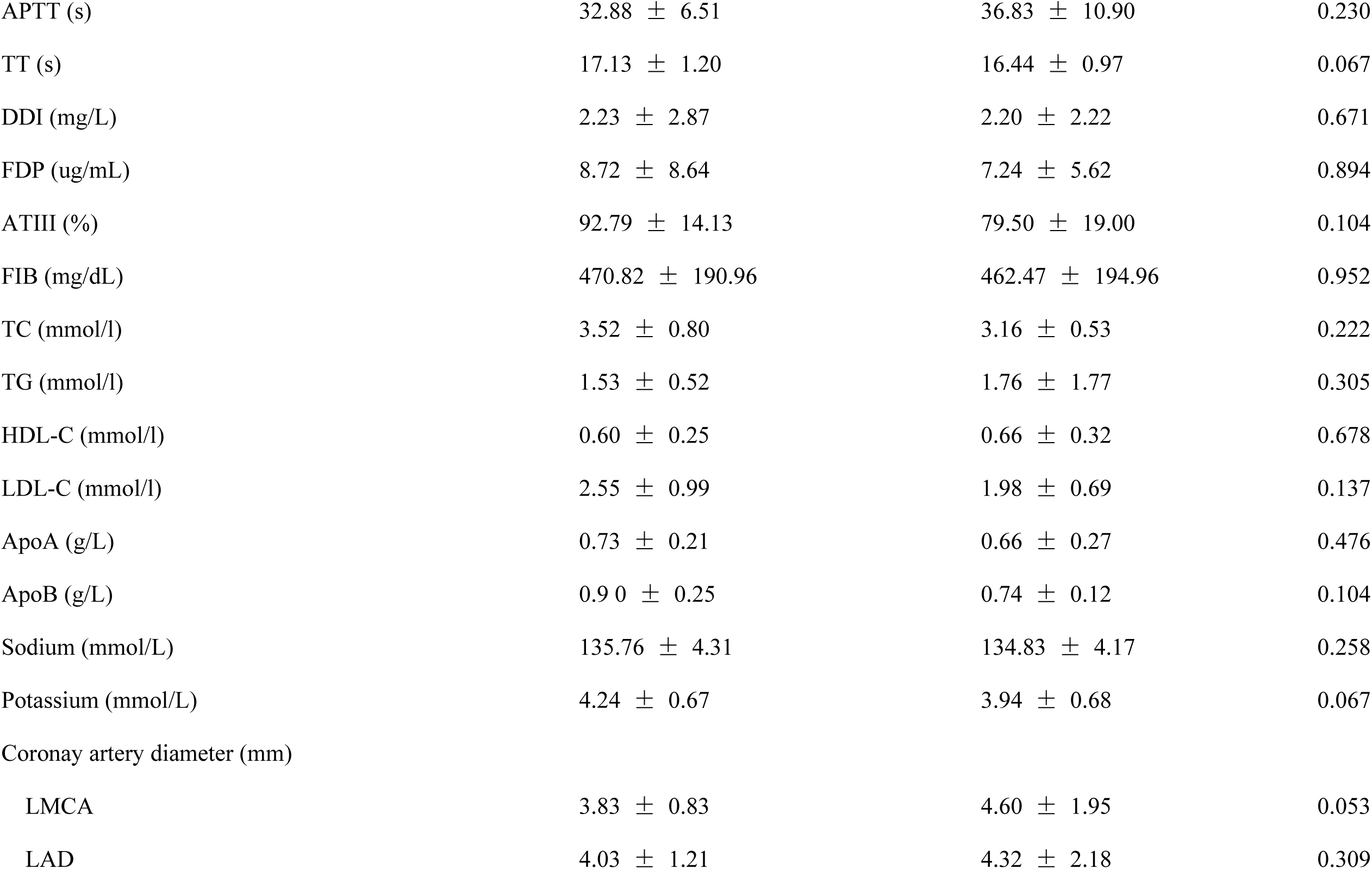
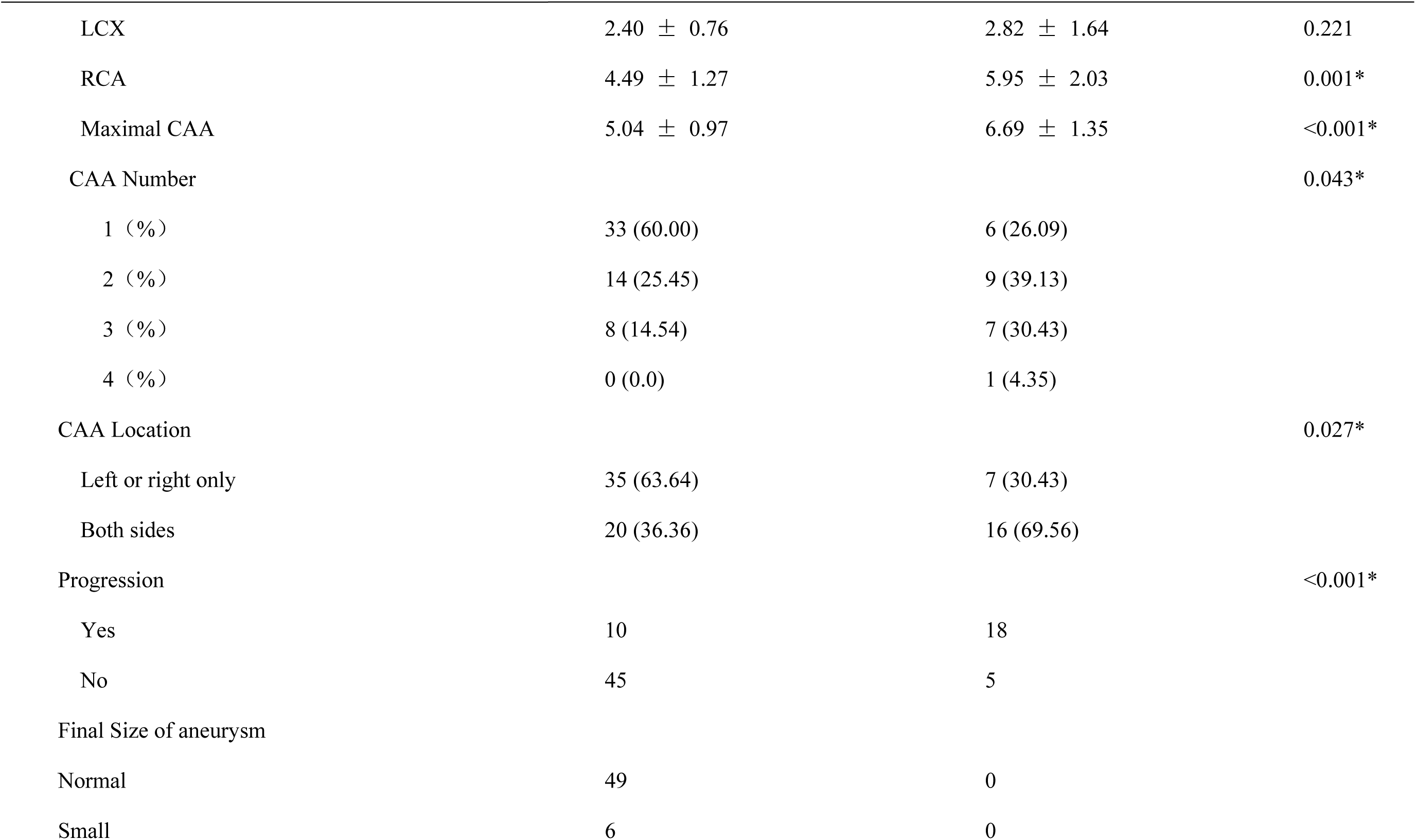
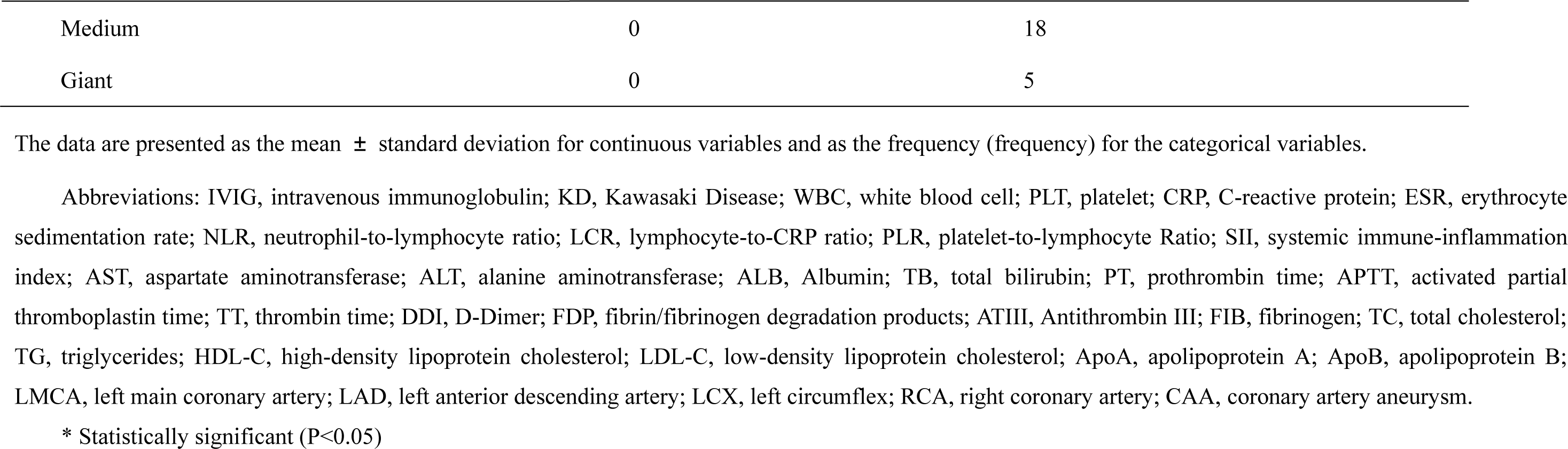
Comparison of Variables Between Regressed Group and Persistent Group in Kawasaki Disease with Medium CAA.

|  | Regressed Group (n=55) | Persistent Group (n=23) | *P* value |
| --- | --- | --- | --- |
| Age(years) | 3.72 ± 2.53 | 4.93 ± 3.19 | 0.131 |
| Male (%) | 39 (68.42) | 16 (76.19) | 0.504 |
| Day of illness before admission, (days) | 7.46 ± 4.01 | 7.06 ± 2.91 | 0.718 |
| Initial IVIG resistance (%) | 19 (34.55) | 9 (42.86) | 0.502 |
| Repeated IVIG resistance (%) | 16 (29.09) | 9 (42.86) | 0.253 |
| Recurrent KD (%) | 1 (1.79) | 3 (14.29) | 0.028* |
| Clinical manifestations |  |  |  |
| Rash, n (%) | 35 (62.50) | 13 (61.90) | 0.962 |
| Bilateral bulbar conjunctive injection, n (%) | 53 (94.64) | 19 (90.48) | 0.509 |
| Edema & erythema of the extremities, n (%) | 25 (44.64) | 10 (47.62) | 0.815 |
| Erythema of oral and pharyngeal mucosa, n (%) | 46 (82.14) | 18 (85.71) | 0.709 |
| Cervical lymphadenopathy, n (%) | 29 (51.79) | 12 (57.14) | 0.675 |
| Incomplete KD, n (%) | 13 (23.21) | 6 (28.57) | 0.627 |
| Pre-IVIG |  |  |  |
| WBC count (10<sup>9</sup>/L) | 15.42 ± 6.66 | 14.54 ± 6.73 | 0.581 |
| Percentage of neutrophil (%) | 66.60 ± 19.68 | 76.29 ± 14.24 | 0.038* |
| Percentage of lymphocyte (%) | 23.62 ± 17.70 | 14.16 ± 8.82 | 0.090 |
| Hemoglobin (g/L) | 109.32 ± 12.31 | 111.14 ± 11.16 | 0.414 |
| PLT count (10<sup>9</sup>/L) | 434.13 ± 219.01 | 351.81 ± 155.33 | 0.288 |
| CRP (mg/L) | 81.21 ± 61.05 | 90.65 ± 39.48 | 0.193 |
| ESR (mm/H) | 81.57 ± 33.56 | 62.47 ± 34.28 | 0.055 |
| NLR | 5.96 ± 5.42 | 7.99 ± 4.74 | 0.079 |
| AST (IU/L) | 64.52 ± 84.05 | 81.25 ± 152.43 | 0.697 |
| ALT (IU/L) | 78.32 ± 140.37 | 96.20 ± 132.53 | 0.294 |
| ALB (g/L) | 37.01 ± 5.46 | 35.43 ± 5.09 | 0.189 |
| Creatinine (umol/L) | 28.66 ± 11.63 | 31.85 ± 15.90 | 0.465 |
| Urea nitrogen (mmol/L) | 3.21 ± 1.39 | 3.53 ± 3.66 | 0.400 |
| TB (umol/L) | 10.62 ± 12.69 | 14.68 ± 20.46 | 0.578 |
| PT (s) | 12.68 ± 2.18 | 15.69 ± 5.53 | 0.014* |
| APTT (s) | 32.88 ± 6.51 | 36.83 ± 10.90 | 0.230 |
| TT (s) | 17.13 ± 1.20 | 16.44 ± 0.97 | 0.067 |
| DDI (mg/L) | 2.23 ± 2.87 | 2.20 ± 2.22 | 0.671 |
| FDP (ug/mL) | 8.72 ± 8.64 | 7.24 ± 5.62 | 0.894 |
| ATIII (%) | 92.79 ± 14.13 | 79.50 ± 19.00 | 0.104 |
| FIB (mg/dL) | 470.82 ± 190.96 | 462.47 ± 194.96 | 0.952 |
| TC (mmol/l) | 3.52 ± 0.80 | 3.16 ± 0.53 | 0.222 |
| TG (mmol/l) | 1.53 ± 0.52 | 1.76 ± 1.77 | 0.305 |
| HDL-C (mmol/l) | 0.60 ± 0.25 | 0.66 ± 0.32 | 0.678 |
| LDL-C (mmol/l) | 2.55 ± 0.99 | 1.98 ± 0.69 | 0.137 |
| ApoA (g/L) | 0.73 ± 0.21 | 0.66 ± 0.27 | 0.476 |
| ApoB (g/L) | 0.90 ± 0.25 | 0.74 ± 0.12 | 0.104 |
| Sodium (mmol/L) | 135.76 ± 4.31 | 134.83 ± 4.17 | 0.258 |
| Potassium (mmol/L) | 4.24 ± 0.67 | 3.94 ± 0.68 | 0.067 |
| Coronay artery diameter (mm) |  |  |  |
| LMCA | 3.83 ± 0.83 | 4.60 ± 1.95 | 0.053 |
| LAD | 4.03 ± 1.21 | 4.32 ± 2.18 | 0.309 |
| LCX | 2.40 ± 0.76 | 2.82 ± 1.64 | 0.221 |
| RCA | 4.49 ± 1.27 | 5.95 ± 2.03 | 0.001* |
| Maximal CAA | 5.04 ± 0.97 | 6.69 ± 1.35 | <0.001* |
| CAA Number |  |  | 0.043* |
| 1 ( % ) | 33 (60.00) | 6 (26.09) |  |
| 2 ( % ) | 14 (25.45) | 9 (39.13) |  |
| 3 ( % ) | 8 (14.54) | 7 (30.43) |  |
| 4 ( % ) | 0 (0.0) | 1 (4.35) |  |
| CAA Location |  |  | 0.027* |
| Left or right only | 35 (63.64) | 7 (30.43) |  |
| Both sides | 20 (36.36) | 16 (69.56) |  |
| Progression |  |  | <0.001* |
| Yes | 10 | 18 |  |
| No | 45 | 5 |  |
| Final Size of aneurysm |  |  |  |
| Normal | 49 | 0 |  |
| Small | 6 | 0 |  |

|  |  |  |
| --- | --- | --- |
| Medium | 0 | 18 |
| Giant | 0 | 5 |

The data are presented as the mean ± standard deviation for continuous variables and as the frequency (frequency) for the categorical variables.
Abbreviations: IVIG, intravenous immunoglobulin; KD, Kawasaki Disease; WBC, white blood cell; PLT, platelet; CRP, C-reactive protein; ESR, erythrocyte sedimentation rate; NLR, neutrophil-to-lymphocyte ratio; LCR, lymphocyte-to-CRP ratio; PLR, platelet-to-lymphocyte Ratio; SII, systemic immune-inflammation index; AST, aspartate aminotransferase; ALT, alanine aminotransferase; ALB, Albumin; TB, total bilirubin; PT, prothrombin time; APTT, activated partial thromboplastin time; TT, thrombin time; DDI, D-Dimer; FDP, fibrin/fibrinogen degradation products; ATIII, Antithrombin III; FIB, fibrinogen; TC, total cholesterol; TG, triglycerides; HDL-C, high-density lipoprotein cholesterol; LDL-C, low-density lipoprotein cholesterol; ApoA, apolipoprotein A; ApoB, apolipoprotein B; LMCA, left main coronary artery; LAD, left anterior descending artery; LCX, left circumflex; RCA, right coronary artery; CAA, coronary artery aneurysm.
\* Statistically significant ( $P < 0.05$ )

### Multivariate analysis analysis and Kaplan-Meier survival

Multivariate logistic regression identified PT and maximal CAA diameter at 1 month as the independent predictors of CAA persistence after adjusting for confounders (Table 2). The 13.6 PT cutoff value was 64.7% sensitive and 73.5% specific [area under the ROC curve (AUC), 0.712, P=0.014]. A 5.65 cutoff value for the maximal CAA at 1month of onset was 85.7% sensitive and 80.7% specific (AUC, 0.857; P<0.001) (Table 3 and Figure 3).

**Figure 3.**
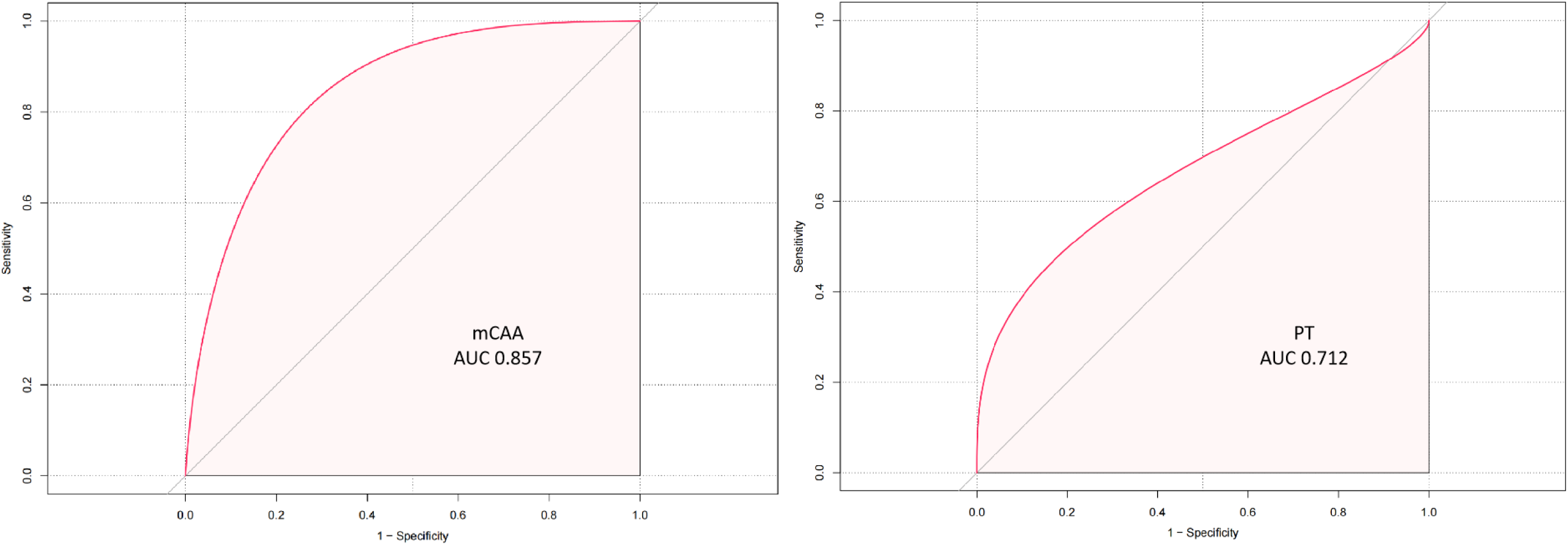
Receiver Operating Characteristic (ROC) curve for maximal CAA and PT in Predicting Persistent Group.

**Table 2.**
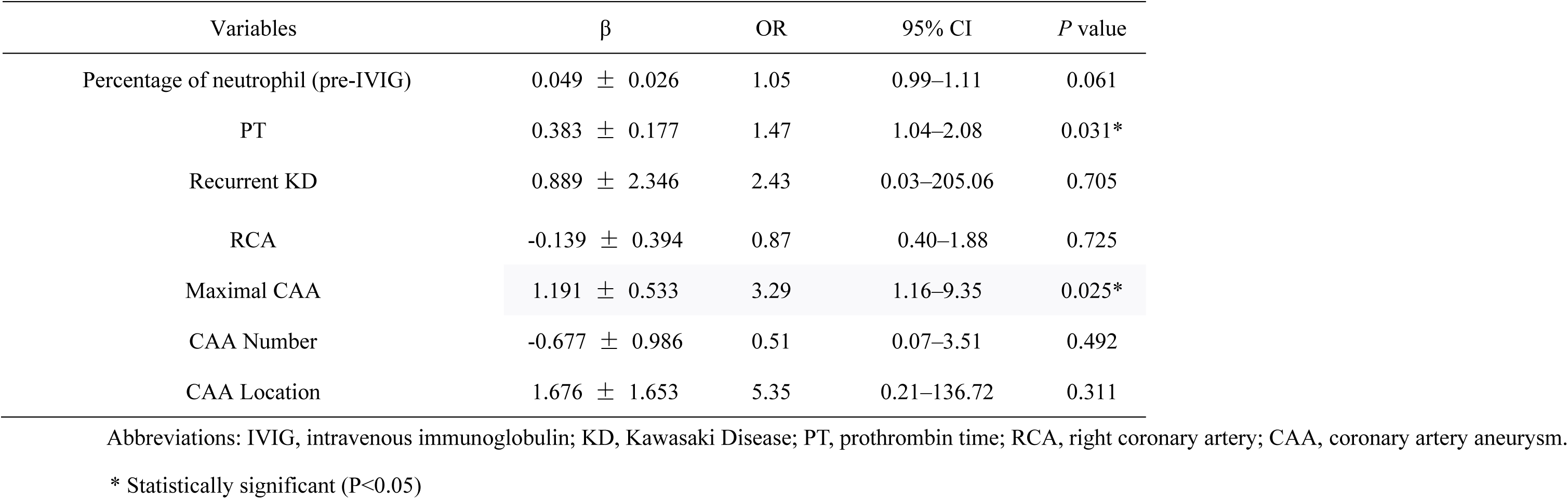
A Multivariate Logistic Regression Model for CAA Persistence in Kawasaki Disease.

| Variables | β | OR | 95% CI | P value |
| --- | --- | --- | --- | --- |
| Percentage of neutrophil (pre-IVIG) | 0.049 ± 0.026 | 1.05 | 0.99–1.11 | 0.061 |
| PT | 0.383 ± 0.177 | 1.47 | 1.04–2.08 | 0.031* |
| Recurrent KD | 0.889 ± 2.346 | 2.43 | 0.03–205.06 | 0.705 |
| RCA | -0.139 ± 0.394 | 0.87 | 0.40–1.88 | 0.725 |
| Maximal CAA | 1.191 ± 0.533 | 3.29 | 1.16–9.35 | 0.025* |
| CAA Number | -0.677 ± 0.986 | 0.51 | 0.07–3.51 | 0.492 |
| CAA Location | 1.676 ± 1.653 | 5.35 | 0.21–136.72 | 0.311 |

Abbreviations: IVIG, intravenous immunoglobulin; KD, Kawasaki Disease; PT, prothrombin time; RCA, right coronary artery; CAA, coronary artery aneurysm.
\* Statistically significant (P<0.05)

**Table 3.** Receiver operating characteristic analysis of PT and maximal CAA for CAA Persistence in Kawasaki Disease.

| Variables | PT | Maximal CAA |
| --- | --- | --- |
| Cut off value | 13.6 | 5.65 |
| Sensitivity % | 64.7 | 85.7 |
| Specificity % | 73.5 | 80.7 |
| AUC | 0.712 | 0.857 |
| P | 0.014* | <0.001* |

Abbreviations: PT, prothrombin time; CAA, coronary artery aneurysm, AUC, area under curve.
\* Statistically significant (P<0.05)

Based on the level of PT, KD patients with medium CAAs were divided into high-PT group (n = 20) and low-PT groups (n = 31) by cut-off of 13.6s. Based on the maximal diameter of CAA at one month, KD patients with medium CAAs were divided into higher group (n = 29) and lower groups (n = 29) by cut-off of 5.6mm. Kaplan-Meier analysis confirmed significant divergence in CAA persistence between groups stratified by these thresholds: PT (log-rank P = 0.0053) and maximal CAA (log-rank P < 0.001) (Figure 4).

**Figure 4.**
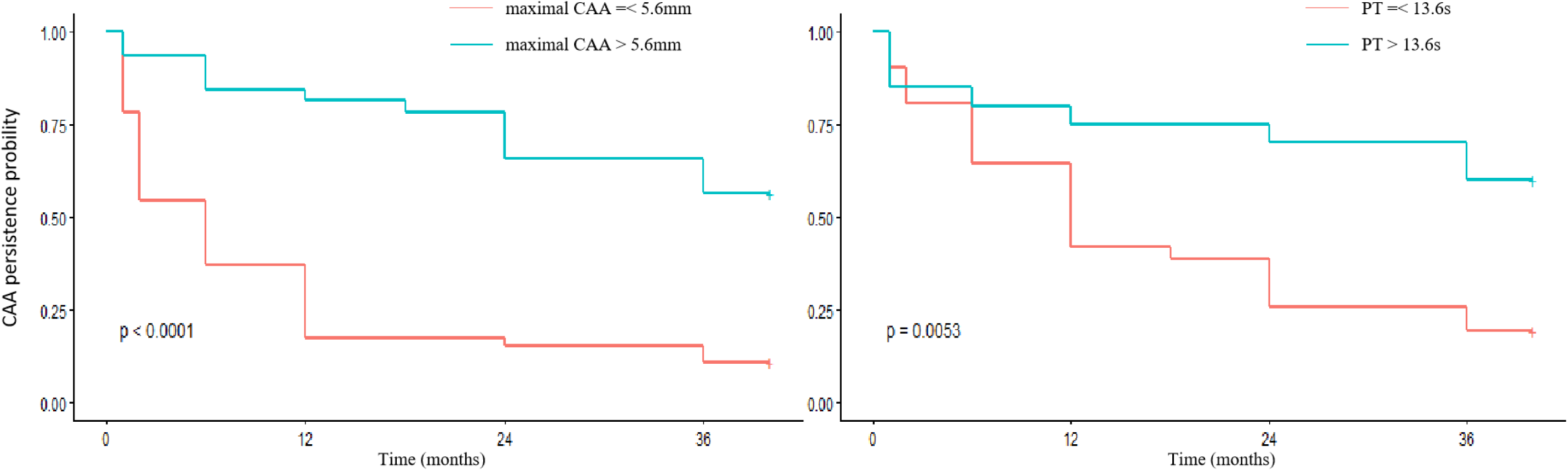
Kaplan-Meier curves for CAA regression.

## Discussion

This prospective study represented the largest reported cohort focusing exclusively on medium CAAs to date. Not only we delineate the natural history of medium CAAs with a follow-up period of 3 years through a longitudinal echocardiography, but predictors of prognostic outcomes in medium CAAs were also explored. We found that the first year represented the period of greatest dynamism in CAA size during follow-up, encompassing both the majority of regressions to small CAA or normal size (78%) and all progression events to giant CAA. Despite this progression, no malignant events such as thrombosis, myocardial infarction, coronary artery stenosis, or death occurred during the follow-up period. This finding highlights the importance of ongoing surveillance and careful management to mitigate potential risks. In addition, we observed that significantly higher pre-IVIG NLR and higher incidence of repeated IVIG resistance as well as lower age of onset might indicate greater chance to progress for patients with medium CAAs. Moreover, we found that the maximal absolutely internal diameter of the coronary artery at 1 month after onset were highly associated with both the progression and persistence of medium-sized CAAs. Importantly, a higher than 13.6s of PT before IVIG refusion during the acute stage possessed predictive power for patients with persistent CAAs. These findings underscore the dual role of morphological severity and systemic inflammation/coagulopathy in driving medium CAA outcomes.

Prior investigations have systematically identified several risk factors or scoring systems to predict CALs in KD patients^20–23^. However, the majority of the included studies enrolled with CAD and small CALs. Later large-scale cohort studies further delineated the natural history of CAAs. For instance, Friedman et al ^4^ analyzed 2860 KD patients (500 with CAAs) in the U.S, identifying progressive risks associated with earlier diagnostic eras (pre-IVIG standardization), baseline medium-giant CAAs, and bilateral involvement. Similarly, Kato et al ^5^ reported that reduced regression rates linked to more than 1 year old at diagnosis, earlier treatment decades, and giant CAAs. Nevertheless, these findings highlight inherent challenges in longitudinal studies, such as evolving protocols (e.g., IVIG timing, adjunctive therapies, monitoring criteria), introducing confounding biases. To address these gaps, our study enrolled a homogeneous cohort under a uniform treatment protocol, controlling for temporal and therapeutic variables, thereby isolating medium CAA-specific risk factors.

Our study observed a higher prevalence of recurrent KD in the persistent CAA group, though multivariate analysis revealed no independent association between recurrence and outcomes. In contrast, prolonged PT consistently emerged as a significant predictor of CAA persistence. This finding aligns with Xi et al’s ^24^ report linking elevated PT to persistent CAL in recurrent KD patients. Notably, our prior work identified PT elevation as a key biomarker in IVIG resistance and Kawasaki disease shock syndrome (KDSS) ^25^, where it synergized with fibrin degradation products, antithrombin III, and D-dimer to predict KDSS onset. Despite conflicting evidence exists, a study reported reduced PT levels in CAL patients^26^, while other cohorts, including Yi et al’s demonstration of PT’s correlation with IVIG resistance^27^ and Yang et al’s identification of PT as an independent predictor of retinol-binding protein 4 (RBP4, a cardiovascular risk marker depleted in CAL patients) ^28^, supported the prognostic relevance of PT.

Mechanistically, prolonged PT may reflect either impaired hepatic synthesis or accelerated consumption of coagulation factors^25^. The liver, a frequent target of KD-related pathology via NK cell-mediated vascular injury, is central to clotting factor production. Concurrently, neutrophil extracellular traps (NETs), pro-thrombotic structures generated by activated neutrophils, exacerbate coagulation cascade activation and complement-driven liver damage, creating a vicious cycle of hepatic dysfunction and hyper-coagulability. This dual pathway underscores the interplay between immune dysregulation, hepatic compromise and coagulopathy in KD progression. Considering the limited predictive ability of single indicator, we can only assume that PT played a role as a surrogate marker integrating multiple pathophysiological processes such as systemic inflammation, impaired liver function, and coagulation abnormalities. These factors collectively contribute to the complex biological mechanisms underlying KD and its complications, including the development and progression of CAAs. Therefore, our finding suggested that CAA progression might not be related simply to the original CAA size but also to the integrative inflammation condition.

CAA diameter, number, and anatomic location have been widely debated as prognostic determinants in KD. While McCrindle et al ^6^ suggested higher regression rates for aneurysms localized in LAD and LCX. Advani et al ^16^ reported no significant association between CAA location and outcomes. Conversely, Zhang et al ^18^ identified multiple CAAs as a predictor of prolonged lesion duration. In our cohort, although CAA multiplicity and location exhibited univariate significance, multivariate analysis revealed maximum CAA diameter at one month as the dominant predictor of persistence, which was consistent with its established role in regression dynamics and cardiovascular risk stratification. Focusing on medium CAAs, we determined an optimal prognostic diameter threshold of 5.65mm, with relatively higher predictive ability, aligning closely with prior studies Chen et al ^29^, who utilized cardiac CT in 18 KD patients, identified 5.6 mm as the critical cutoff, while Tsuda et al ^17^ derived a similar threshold of 5.7 mm through longitudinal analysis of a total of 195 CAAs. This remarkable consistency across methodologies suggests that a diameter of ∼5.6–5.7 mm may demarcate a hemodynamic tipping point, beyond which endothelial shear stress and impairing vascular repair mechanisms.

The temporal trajectory of CAA evolution has been well-characterized. Tsuda et al ^17^ reported median CAA emerged at 12 days post-onset, peaking around day 30, while parallel study ^30^ noted progressive dilatation rates escalating through day 10. These observations aligned with the established timeline of acute vasculitis, which persists for up to seven weeks after disease onset ^31^. During this critical period, the systemic inflammation of KD leads to widespread vascular damage, particularly affecting the coronary arteries, supporting the inflammatory process is a key driver of CAA formation and progression. Our findings further implicated repeated IVIG resistance, elevated post-IVIG neutrophil percentage, and maximal CAA diameter at one month as predictors of CAA progression (data not shown), which directly reflected the intensity of acute-phase inflammation and structural compromise. These factors exacerbate the inflammatory response and prolong vascular damage, increasing the likelihood of aneurysm development. Similarly, the timeline of acute vasculitis provides a framework for understanding the temporal relationship between laboratory abnormalities and clinical outcomes in KD. Collectively, these associations suggest that CAA progression may signify persistent sub-clinical inflammation or hemodynamic abnormality, perpetuating a cycle of vascular remodeling. This hypothesis warrants validation through the targeted studies integrating serial biomarkers and advanced imaging modalities.

### Limitations

This study has several potential limitations. Firstly, our hospital is the largest pediatric medical center in Southwest China, which may lead to a selection bias due to a higher number of severely ill patients being admitted to this facility. Secondly, our study was performed with strict inclusion and exclusion criteria. The findings of this study are, therefore, applicable only to Chinese KD patients receiving the standardized IVIG treatment (2 g/kg) within 10 days of the onset of fever. Thirdly, the follow-up duration was limited to three years, extending this period may be warranted in future research.Lastly, despite sourcing cases from a high-volume pediatric referral center over a 9-year span, the rarity of KD with medium CAA yielded a limited sample size, reducing analytical robustness and precision. Although the current sample size is limited, this cohort represents the largest reported prospective series specifically focused on medium CAAs to date. Future investigations should prioritize multi-institutional collaborations to assemble larger cohorts for validation.

## Conclusion

Patients with medium CAAs who have higher PT of pre-IVIG during the acute stage and maximal diameters of CAA at 1month of onset were more likely to develop large-sized CAA. Nevertheless, progressive coronary dilatation may increase the risk of more severe coronary damage and dramatically prolong coronary artery normalization time. Thus, aggressive risk modifications should be designed for the surveillance and prevention of late cardiovascular events.

## Data Availability

All data referred to in the manuscript will be available.

## Funding support and author disclosures

This work was supported by the Natural Science Foundation of Sichuan Province (2024YFFK0272); the Key Research and Development Project of Chengdu Science and Technology Bureau (2024-YF05-00237-SN); the National Natural Science Foundation of China (No. 82370236); and the National Key Research and Development Program of China (No. 2022YFC2703902). The authors read and approved the final manuscript and declare no competing interests.

---

**Supplementary Table 1.** Comparison of Post-IVIG Laboratory Variables Between Regressed Group and Persistent Group in Kawasaki Disease with Medium CAA.

|  | Regressed Group (n=55) | Persistent Group (n=23) | *P* value |
| --- | --- | --- | --- |
| WBC count ($10^9/L$) | 10.21 ± 6.82 | 10.68 ± 5.43 | 0.461 |
| Percentage of neutrophil (%) | 48.59 ± 20.00 | 57.32 ± 23.87 | 0.122 |
| Percentage of lymphocyte (%) | 39.97 ± 19.73 | 31.04 ± 19.88 | 0.115 |
| Hemoglobin (g/L) | 107.19 ± 11.55 | 104.10 ± 10.90 | 0.338 |
| PLT count ($10^9/L$) | 528.54 ± 235.09 | 430.38 ± 153.94 | 0.149 |
| CRP (mg/L) | 26.26 ± 26.51 | 43.34 ± 40.42 | 0.085 |
| NLR | 2.12 ± 2.38 | 3.75 ± 3.79 | 0.121 |

The data are presented as the mean ± standard deviation for continuous variables and as the frequency (frequency) for the categorical variables.
Abbreviations: IVIG, intravenous immunoglobulin; WBC, white blood cell; PLT, platelet; CRP, C-reactive protein; ESR, erythrocyte sedimentation rate; NLR, neutrophil-to-lymphocyte ratio; LCR, lymphocyte-to-CRP ratio; PLR, platelet-to-lymphocyte Ratio; SII, systemic immune-inflammation index.
\* Statistically significant (P<0.05)

## Notes

### Competing Interest Statement

The authors have declared no competing interest.

### Author Declarations

The University Ethics Committee on Human Subjects at Sichuan University

## References

1. Fukazawa R, Kobayashi J, Ayusawa M, Hamada H, Miura M, Mitani Y, Tsuda E, Nakajima H, Matsuura H, Ikeda K, Nishigaki K, Suzuki H, Takahashi K, Suda K, Kamiyama H, Onouchi Y, Kobayashi T, Yokoi H, Sakamoto K, Ochi M, Kitamura S, Hamaoka K, Senzaki H and Kimura T. JCS/JSCS 2020 Guideline on Diagnosis and Management of Cardiovascular Sequelae in Kawasaki Disease. Circulation Journal. 2020;84:1348–1407.

2. McCrindle BW, Rowley AH, Newburger JW, Burns JC, Bolger AF, Gewitz M, Baker AL, Jackson MA, Takahashi M, Shah PB, Kobayashi T, Wu M-H, Saji TT and Pahl E. Diagnosis, Treatment, and Long-Term Management of Kawasaki Disease: A Scientific Statement for Health Professionals From the American Heart Association. Circulation. 2017;135.

3. Jone P-N, Tremoulet A, Choueiter N, Dominguez SR, Harahsheh AS, Mitani Y, Zimmerman M, Lin M-T and Friedman KG. Update on Diagnosis and Management of Kawasaki Disease: A Scientific Statement From the American Heart Association. Circulation. 2024;150.

4. Friedman KG, Gauvreau K, Hamaoka-Okamoto A, Tang A, Berry E, Tremoulet AH, Mahavadi VS, Baker A, deFerranti SD, Fulton DR, Burns JC and Newburger JW. Coronary Artery Aneurysms in Kawasaki Disease: Risk Factors for Progressive Disease and Adverse Cardiac Events in the US Population. Journal of the American Heart Association. 2016;5.

5. Kato T, Miura M, Kobayashi T, Kaneko T, Fukushima N, Suda K, Maeda J, Shimoyama S, Shiono J, Hirono K, Ikeda K, Sato S, Numano F, Mitani Y, Waki K, Ayusawa M, Fukazawa R, Fuse S, Miura M, Fukazawa R, Fuse S, Hamaoka K, Hirono K, Kato T, Kato H, Kobayashi T, Saji T, Suda K, Waki K, Yamagishi H, Fukushima N, Tomotsune M, Yoshida M, Kaneko T, Toyono M, Furuno K, Shimoyama S, Iwashima S, Moritou Y, Kamada M, Takeda A, Shiono J, Sano T, Omori D, Fukasawa Y, Mii S, Nomura Y, Nakamura T, Maeda J, Ishii M, Ogata S, Kitagawa A, Yamamoto M, Ikeda K, Yamamura K, Mitani Y, Masuda H, Kaneko M, Kawamura Y, Komori A, Ayusawa M, Sato S, Numano F, Suzuki H, Watanabe K, Hayashi M, Watanabe M, Kuraishi K, Nishihara E, Katayama H, Okumura K, Takahashi T, Horita N, Matsuzaki S, Motoki N, Akazawa Y, Aso K, Nagumo K, Takatuki S, Suganuma E, Matsuda S, Hayabuchi Y, Doi S, Honda T, Terai M and Miyamoto T. Analysis of Coronary Arterial Aneurysm Regression in Patients With Kawasaki Disease by Aneurysm Severity: Factors Associated With Regression. Journal of the American Heart Association. 2023;12.

6. McCrindle BW, Manlhiot C, Newburger JW, Harahsheh AS, Giglia TM, Dallaire F, Friedman K, Low T, Runeckles K, Mathew M, Mackie AS, Choueiter NF, Jone PN, Kutty S, Yetman AT, Raghuveer G, Pahl E, Norozi K, McHugh KE, Li JS, De Ferranti SD, Dahdah N, Altman CA, Anderson BR, Beaulieu E, Boychuk CE, Braunlin E, Burns JC, Carr MR, Crean A, Colyer JH, Dempsey A, Desjardins L, Dillenburg R, Dionne A, Ferris A, Gewitz M, Grcic MM, Greenway SC, Harris KC, Hayden-Rush C, Hill KD, Jain S, Kimball TR, Lang SM, Lin MT, Mahle WT, Mondal T, Portman MA, Renaud C, Sexson Tejitel SK, Szmuszkovicz JR, Texter KM, Thacker D, Tierney ESS, Thomas T, Tremoulet AH, Wagner-Lees S and Warren A. Medium-Term Complications Associated With Coronary Artery Aneurysms After Kawasaki Disease: A Study From the International Kawasaki Disease Registry. Journal of the American Heart Association. 2020;9.

7. Miura M, Kobayashi T, Kaneko T, Ayusawa M, Fukazawa R, Fukushima N, Fuse S, Hamaoka K, Hirono K, Kato T, Mitani Y, Sato S, Shimoyama S, Shiono J, Suda K, Suzuki H, Maeda J, Waki K, Kato H, Saji T, Yamagishi H, Ozeki A, Tomotsune M, Yoshida M, Akazawa Y, Aso K, Doi S, Fukasawa Y, Furuno K, Hayabuchi Y, Hayashi M, Honda T, Horita N, Ikeda K, Ishii M, Iwashima S, Kamada M, Kaneko M, Katyama H, Kawamura Y, Kitagawa A, Komori A, Kuraishi K, Masuda H, Matsuda S, Matsuzaki S, Mii S, Miyamoto T, Moritou Y, Motoki N, Nagumo K, Nakamura T, Nishihara E, Nomura Y, Ogata S, Ohashi H, Okumura K, Omori D, Sano T, Suganuma E, Takahashi T, Takatsuki S, Takeda A, Terai M, Toyono M, Watanabe K, Watanabe M, Yamamoto M and Yamamura K. Association of Severity of Coronary Artery Aneurysms in Patients With Kawasaki Disease and Risk of Later Coronary Events. JAMA Pediatrics. 2018;172.

8. Tsuda E, Kamiya T, Ono Y, Kimura K, Kurosaki K and Echigo S. Incidence of Stenotic Lesions Predicted by Acute Phase Changes in Coronary Arterial Diameter During Kawasaki Disease. Pediatric Cardiology. 2004;26:73–79.

9. Tsuda E, Tsujii N and Hayama Y. Stenotic Lesions and the Maximum Diameter of Coronary Artery Aneurysms in Kawasaki Disease. The Journal of Pediatrics. 2018;194:165–170.e2.

10. Tsuda E, Hamaoka K, Suzuki H, Sakazaki H, Murakami Y, Nakagawa M, Takasugi H and Yoshibayashi M. A survey of the 3-decade outcome for patients with giant aneurysms caused by Kawasaki disease. American Heart Journal. 2014;167:249–258.

11. Peng Y and Yi Q. Incidence and timing of coronary thrombosis in Kawasaki disease patients with giant coronary artery aneurysm. Thrombosis Research. 2023;221:30–34.

12. Elias MD, Brothers JA, Hogarty AN, Martino J, O’Byrne ML, Patel C, Stephens P, Tingo J, Vetter VL, Ravishankar C and Giglia TM. Outcomes Associated with Giant Coronary Artery Aneurysms after Kawasaki Disease: A Single-Center United States Experience. The Journal of Pediatrics. 2024;274.

13. Dietz SM, Kuipers IM, Koole JCD, Breur JMPJ, Fejzic Z, Frerich S, Dalinghaus M, Roest AAW, Hutten BA and Kuijpers TW. Regression and Complications of z-score-Based Giant Aneurysms in a Dutch Cohort of Kawasaki Disease Patients. Pediatric Cardiology. 2017;38:833–839.

14. Liu L, Luo C, Hua Y, Wu M, Shao S, Liu X, Zhou K and Wang C. Risk factors associated with progression and persistence of small- and medium-sized coronary artery aneurysms in Kawasaki disease: a prospective cohort study. European Journal of Pediatrics. 2020;179:891–900.

15. Liu J, Yue Q, Qin S, Su D, Ye B and Pang Y. Risk factors and coronary artery outcomes of coronary artery aneurysms differing in size and emergence time in children with Kawasaki disease. Frontiers in Cardiovascular Medicine. 2022;9.

16. Advani N, Sastroasmoro S, Ontoseno T and Uiterwaal C. Long-term outcome of coronary artery dilatation in Kawasaki disease. Annals of Pediatric Cardiology. 2018;11.

17. Tsuda E and Hashimoto S. Time Course of Coronary Artery Aneurysms in Kawasaki Disease. The Journal of Pediatrics. 2021;230:133–139.e2.

18. Zhang X, He Y, Shao Y, Hang B, Xu Z and Chu M. Factors affecting the duration of coronary artery lesions in patients with the Kawasaki disease: a retrospective cohort study. Pediatric Rheumatology. 2021;19.

19. Chih W-L, Wu P-Y, Sun L-C, Lin M-T, Wang J-K and Wu M-H. Progressive Coronary Dilatation Predicts Worse Outcome in Kawasaki Disease. The Journal of Pediatrics. 2016;171:78–82.e1.

20. Ae R, Maddox RA, Abrams JY, Schonberger LB, Nakamura Y, Kuwabara M, Makino N, Kosami K, Matsubara Y, Matsubara D, Sasahara T and Belay ED. Kawasaki Disease With Coronary Artery Lesions Detected at Initial Echocardiography. Journal of the American Heart Association. 2021;10.

21. Hua W, Ma F, Wang Y, Fu S, Wang W, Xie C, Zhang Y and Gong F. A new scoring system to predict Kawasaki disease with coronary artery lesions. Clinical Rheumatology. 2018;38:1099–1107.

22. Xie L-p, Yan W-l, Huang M, Huang M-r, Chen S, Huang G-y and Liu F. Epidemiologic Features of Kawasaki Disease in Shanghai From 2013 Through 2017. Journal of Epidemiology. 2020;30:429–435.

23. Zhao L, Wu J, Liu X, Zhou K, Hua Y, Shao S and Wang C. Risk factors for predicting medium-giant coronary artery aneurysms in Kawasaki disease. Immunologic Research. 2025;73.

24. Chen X, Gao L, Zhen Z, Wang Y, Na J, Yu W, Tian Z, Yuan Y and Qian S. Incidence of coronary artery lesions in children with recurrent Kawasaki disease. Expert Review of Clinical Immunology. 2024;20:673–678.

25. Li B, Liu X, Shao S, Wu P, Wu M, Liu L, Hua Y, Duan H, Zhou K and Wang C. Predictive value of coagulation profiles for Kawasaki disease shock syndrome: a prospective cohort study. Frontiers in Pediatrics. 2024;12.

26. Yin Q-G, Zhou J, Zhou Q, Shen L, Zhang M-Y and Wu Y-H. Diagnostic performances of D-dimer, prothrombin time, and red blood cell distribution width for coronary artery lesion in children with acute stage Kawasaki disease. Frontiers in Pediatrics. 2023;11.

27. Dae Yong Yi JYK, Eun Young Choi, Jung Yun Choi and Hye Ran Yang. Hepatobiliary risk factors for clinical outcome of Kawasaki disease in children. BMC Pediatrics 2014.

28. Yang M, Weng H, Pei Q, Jing F, Yi Q and Andrukhov O. The Relationship between Retinol-Binding Protein 4 and Markers of Inflammation and Thrombogenesis in Children with Kawasaki Disease. Mediators of Inflammation. 2021;2021:1–7.

29. Chen P-T, Lin M-T, Chen Y-S, Chen S-J and Wu M-H. Computed tomography predict regression of coronary artery aneurysm in patients with Kawasaki disease. Journal of the Formosan Medical Association. 2017;116:806–814.

30. Fuse S, Mori T, Kuroiwa Y and Hirakawa S. On What Day of Illness Does the Dilatation of Coronary Arteries in Patients With Kawasaki Disease Begin? Circulation Journal. 2018;82:247–250.

31. Naoe S, Takahashi K, Masuda H and Tanaka N. Kawasaki Disease With Particular Emphasis on Arterial Lesions. Acta Pathologica Japonica. 2008;41:785–797.

